# Comparing Non-Communicable Disease Risk Factors in Asian Migrants and Native Koreans

**DOI:** 10.1101/2020.12.22.20248150

**Authors:** Heng Piao, Jae Moon Yun, Aesun Shin, Daehee Kang, Belong Cho

## Abstract

**Importance:** Regarding international migrants, the theories of healthy migration effect and sick migration effect both exist; thus, assessing the health of international migrants is crucial in the Republic of Korea, Asia, and even worldwide.

**Objective:** To compare non-communicable disease risk factors among Asian migrants in Korea and the Korean population.

**Design:** A cross-sectional (2015) and longitudinal (2009□2015) observational study.

**Setting:** Population-wide analysis using the National Health Information Database of the Korean National Health Insurance Service for 2009□2015.

**Participants:** Asian migrants (n=987,214) in Korea and Korean nationals (n=1,693,281) aged ≥20 years were included. In addition, Asian migrants were divided into Chinese, Japanese, Filipino, Vietnamese, and other Asian migrants.

**Exposure:** The nationality of Asian migrants compared with Koreans.

**Main Outcomes and Measures:** The prevalence of non-communicable disease risk factors, such as current smoking, obesity, diabetes, and hypertension, in 2015 was analyzed. Regarding the age-adjusted prevalence, direct age standardization was conducted separately by sex using 10-year age bands; the World Standard Population was used as the standard population.

**Results:** Among participants aged ≥20 years, the age-adjusted prevalence of current smoking was higher among Chinese migrant men than among Korean men (P<0.001) and among other Asian migrant women than among Korean women (P<0.001). The age-adjusted prevalence of obesity was higher in Chinese, Filipino, and other Asian migrant women than in Korean women (P<0.001, P=0.002, and P<0.001, respectively). Among participants aged 20–49 years, the age-adjusted prevalence of diabetes and hypertension was higher in Filipino migrant women than in Korean women (P=0.009 and P<0.001, respectively).

**Conclusion and Relevance:** The current status of smoking and obesity among Asian migrants of specific nationalities is worse than that among native Koreans. Moreover, the health inequalities among Filipino migrant women in Korea, especially those aged 20–49 years, should be addressed.

**Key Points:** *Question:* Do international migrants in Korea have a health advantage regarding non-communicable disease risk factors?

*Findings:* Among participants aged ≥20 years, the problem of current smoking among Chinese migrant men and other Asian migrant women and that of obesity among Chinese, Filipino, and other Asian migrant women in Korea needs to be addressed. The prevalence of diabetes and hypertension was higher among Filipino migrant women than among Korean women in the 20–49-year age group.

*Meaning:* The relationships between Asian migrant nationality and non-communicable disease risk factors provide evidence for targeting high-risk groups and improving policy development in Korea.

## Introduction

Recent systematic reviews and meta-analyses of migrant health have supported the healthy migration hypothesis that international migrants have health advantages.^1,2^ This healthy migration effect suggests that international migrants, particularly new ones, are healthier than the overall host country population^1^; this can be attributed to self-selection or immigration policies^1,3^ but is inversely associated with the duration of residence in the host country.^1,4^ However, in Israel, a sick migration effect is shown among international migrants within the initial 20 years of residence since commencement, implying that international migrants have health disadvantages compared with the native population in the host country.^5^

Compared with other ethnic groups, South Asians are more likely to have diabetes and cardiovascular disease (CVD), possibly due to socioeconomic influences, the residence region, and migration.^6^ The South Asian migrant population has a higher risk of diabetes than the European population, and onset is 5–10 years earlier, despite lower body mass indices (BMIs).^7^ In the Republic of Korea, compared with Korean natives, the prevalence of obesity in Asian migrants showed increasing trends, with it becoming higher in Asian migrant women than in Korean native women since 2014.^8^

Few previous studies have involved Asian migrants (those born in Asian countries and moving to another high-income East Asian country) using nationwide data.^2,4,7^ Therefore, we investigated differences in the prevalence of non-communicable disease (NCD) risk factors between Asian migrants of different nationalities and the Korean population to evaluate whether there is a healthy migration effect in Korea using high-quality health check-up data from the National Health Insurance Service (NHIS). Moreover, we investigated differences in obesity, diabetes, and elevated blood pressure among Chinese, Filipino, and Vietnamese migrants in Korea compared with the general population of their home countries. Based on our previous study,^8^ we hypothesized that Asian migrants of different nationalities have greater health advantages regarding NCD risk factors than Koreans.

## Methods

### Study Population

We conducted an observational retrospective population-based analysis of data collected during a longitudinal study. The study population constituted Asian migrants and the general Korean population aged ≥20 years who underwent health check-ups between 2009 and 2015; data were obtained from the National Health Information Database (NHID) established by the Korean NHIS.^8,9^ The NHID included eligibility data, national health check-up data, healthcare utilization, long-term care insurance, and health care provider information.^9^ Information on income-based insurance contributions, demographic variables, and date of death was included.^9^ The NHID is suitable for population-based studies aimed at primary and secondary prevention of diseases.^9,10^ The health check-up data for Asian migrants could be extracted from customized NHID data; this data represented most Asian migrants who underwent annual voluntary health check-ups in Korea.^9,11^ The health check-up data of the Korean population were obtained from the 1 million-sample cohort of the NHID, a random sample representing 2.2% of the Korean-born population, representative of the nationwide Korean population.^12^ Considering that the NHID is more extensive than it seems, the NHIS conducted systematic stratified random sampling with 2.2% within each stratum for the construction of the 1 million-sample cohort, of which the target variable for sampling was the individual’s total annual medical expenses.^12^

Asian migrants originated from China, Japan, Vietnam, the Philippines, Indonesia, Thailand, Uzbekistan, Sri Lanka, Mongolia, Bangladesh, Pakistan, and India. They were divided into Chinese, Japanese, Filipino, Vietnamese, and other Asian migrants to ensure sufficient samples after age standardization in 2015 (eMethods).

### Measurements

Age, sex, and monthly insurance premiums are included in the data collected by the NHIS; insurance premiums are determined by income level in Korea.^8,13^ Prevalence was defined as the number of participants with cases in a given year per number of participants in the population who underwent a health check-up during this period. Incidence was defined as the number of participants with new cases during the specified period per number of participants in the population who underwent health check-ups more than once during this period (eMethods).

The primary outcome was the prevalence of NCD risk factors, including current smoking, physical inactivity, obesity, diabetes, elevated blood pressure, hypertension, and hypercholesterolemia, in 2015.^14–16^ The secondary outcomes were the incidence of type 2 diabetes (T2D) and hypertension during 2009–2015. To ascertain incident T2D and hypertension, we excluded participants with diabetes and hypertension at the first health check-up between 2009 and 2014. Incident T2D and hypertension were determined for each participant between the first and last health check-ups.^11^

### IRB Approval

The Institutional Review Board (IRB) of Seoul National University Hospital approved the study on August 21, 2018 (IRB number: E-1808-093-966) and waived the need for participant informed consent.

### Statistics

We conducted several sets of analyses. The research design used and statistical analyses performed for this and previous studies are shown in eFigure 1. First, to calculate the age-adjusted prevalence of lifestyle, socioeconomic, and health-related factors among Chinese, Japanese, Filipino, Vietnamese, and other Asian migrants compared with Koreans aged ≥20 years, direct age standardization was conducted by sex using 10-year age bands, wherein the World Standard Population was used as the standard population.^17^ For Asian migrants compared with Koreans aged 20–29, 30–39, and 40–49 years, we examined age-adjusted prevalence ratios (aPRs) and 95% log-normal confidence intervals (CIs).^18^ Moreover, we conducted direct age standardization in Asian migrants compared with Koreans aged 20–49 years, wherein the World Standard Population was used as the standard population.^17^ Demographic and risk factor variables were compared between Asian migrants and Koreans, according to nationality, using unpaired 2-tailed t tests for continuous variables and χ^2^ tests for categorical variables.

Second, to examine the differences in the development of T2D and hypertension between Asian migrants according to nationality and the native Korean population, multivariable logistic regression analyses were conducted and adjusted for the following covariates: age (continuous, years), sex, economic status, BMI (continuous, kg/m^2^), smoking status, alcohol use, and physical activity. The adjusted odds ratios (ORs) and 95% CIs for incident T2D and hypertension determinants were examined between the first and last health check-ups during 2009–2015. Further, we conducted multivariable logistic regression analyses stratified by age (20–39 and ≥40 years) and sex.^11^

Finally, we examined differences in the age-adjusted prevalence of obesity, elevated blood pressure, and diabetes among Chinese, Filipino, and Vietnamese migrants compared with the general population of their respective home countries.^11^ Age standardization was conducted using 10-year age bands, wherein the World Standard Population was used as the standard population.^17^ Health data among general populations in China, the Philippines, and Vietnam were obtained from the Global Health Observatory Data Repository of the World Health Organization (available from: http://apps.who.int/gho/data/node.main.A867?lang=en/) (eMethods).

All analyses were performed using SAS version 9.3 (SAS Institute Inc., Cary, NC, USA). P-values <0.05 were considered statistically significant.

## Results

Table 1 presents the characteristics of Asian migrants of different nationalities and Koreans in 2015.The age-adjusted prevalence of current smoking in 2015 was higher in Chinese migrant men than in Korean men (P<0.001; Table 2) and higher in other Asian migrant women than in Korean women (P<0.001; Table 3). The age-adjusted prevalence of obesity was higher in Chinese, Filipino, and other Asian migrant women than in Korean women (P<0.001, P=0.002, and P<0.001, respectively; Table 3). The age-adjusted prevalence of hypercholesterolemia was higher in Japanese and Vietnamese migrant men than in Korean men (P=0.004 and P=0.016, respectively; Table 2) and higher in Japanese and Vietnamese migrant women than in Korean women (P<0.001 and P<0.001, respectively; Table 3).

**Table 1.**
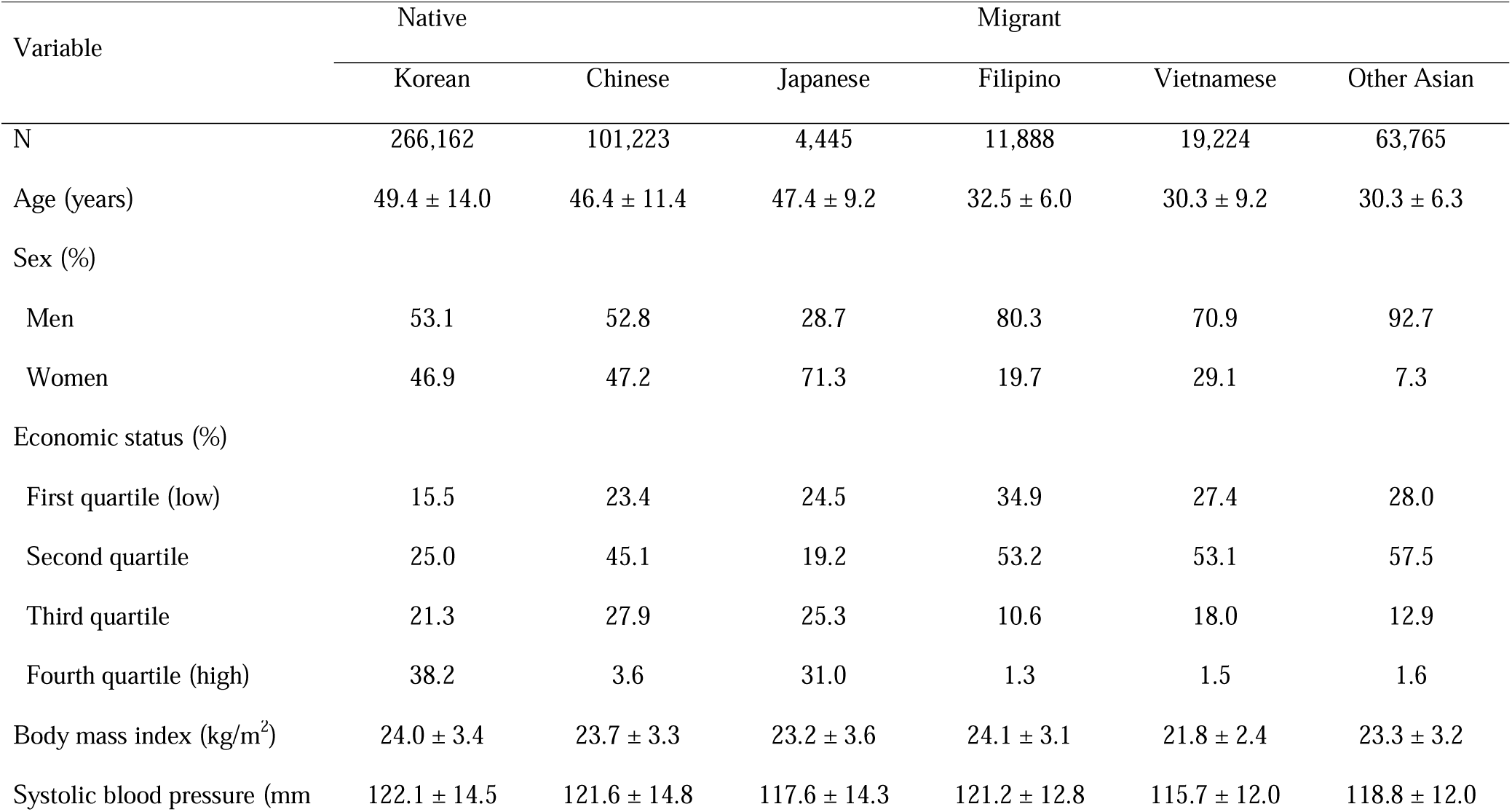
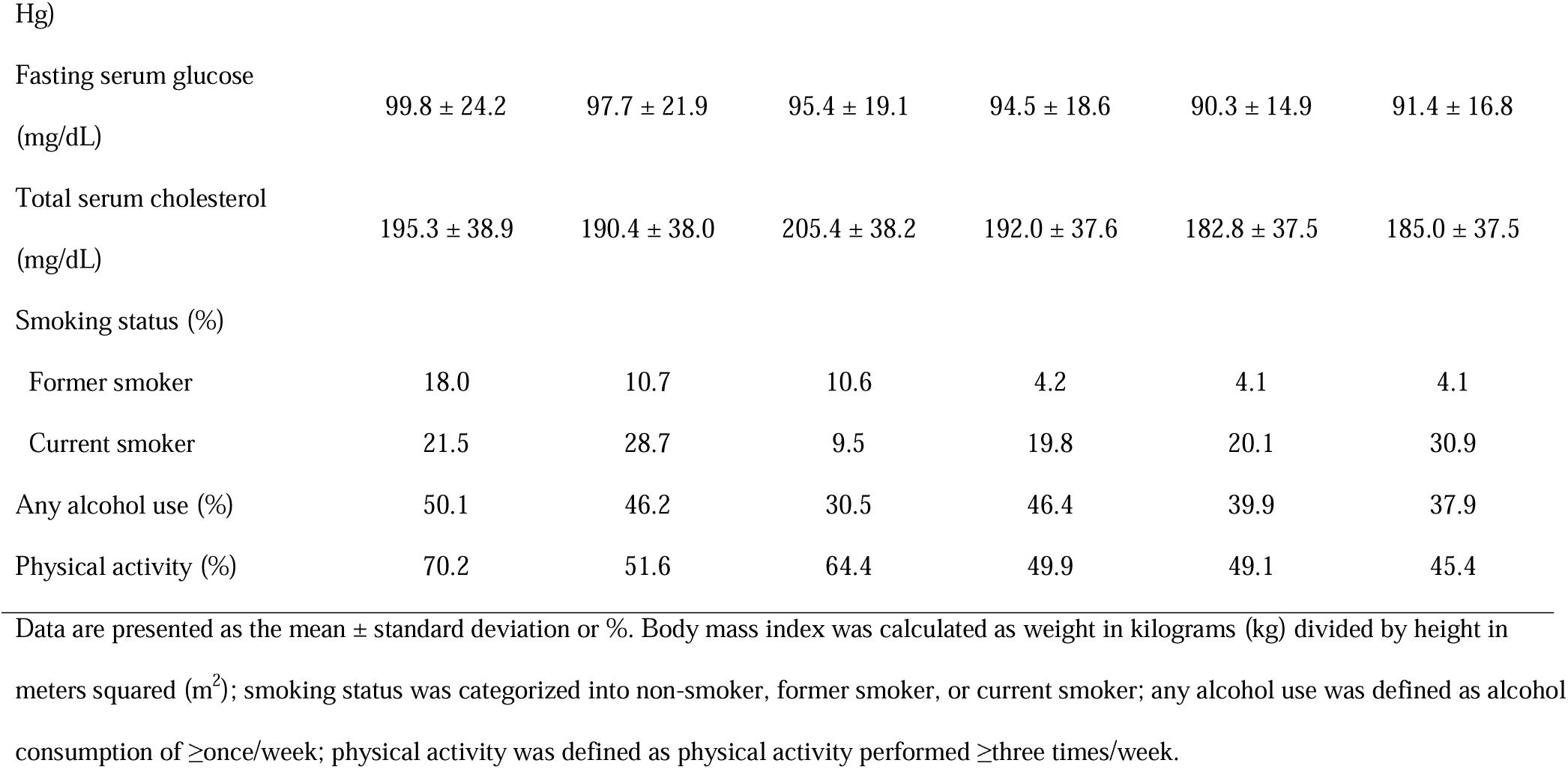
Health characteristics of Asian migrants and Koreans aged ≥20 years in 2015

**Table 2.**
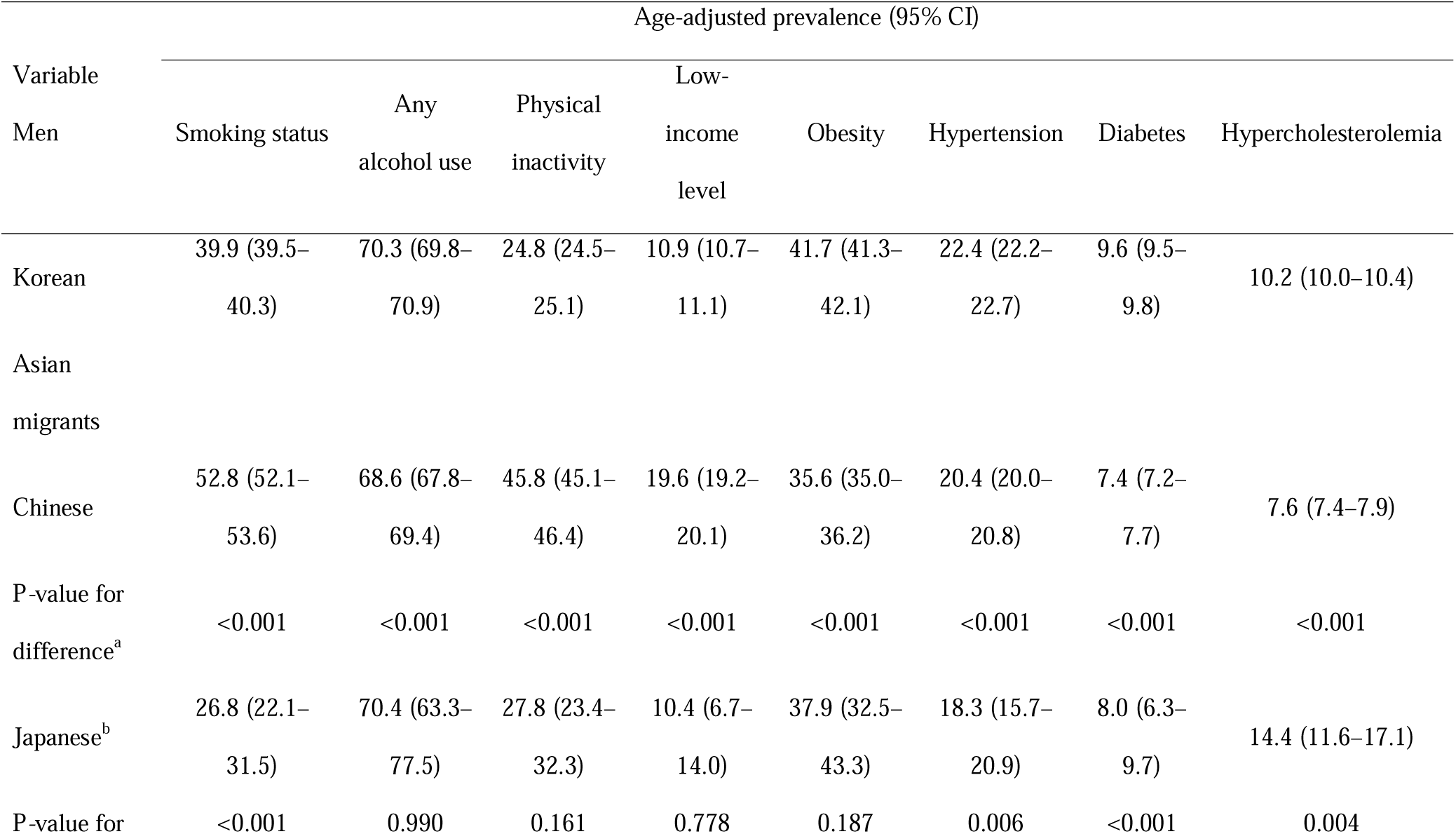
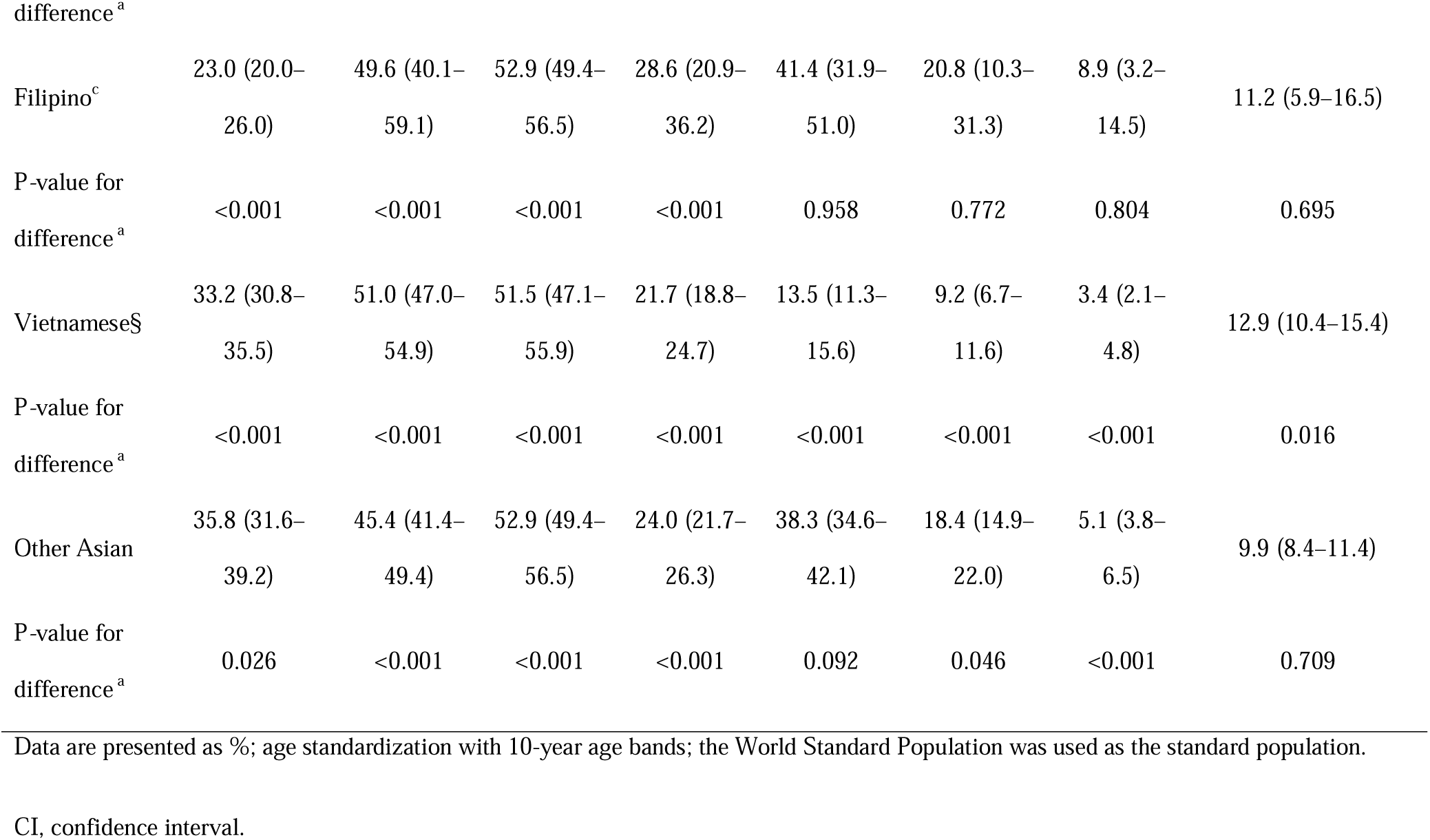
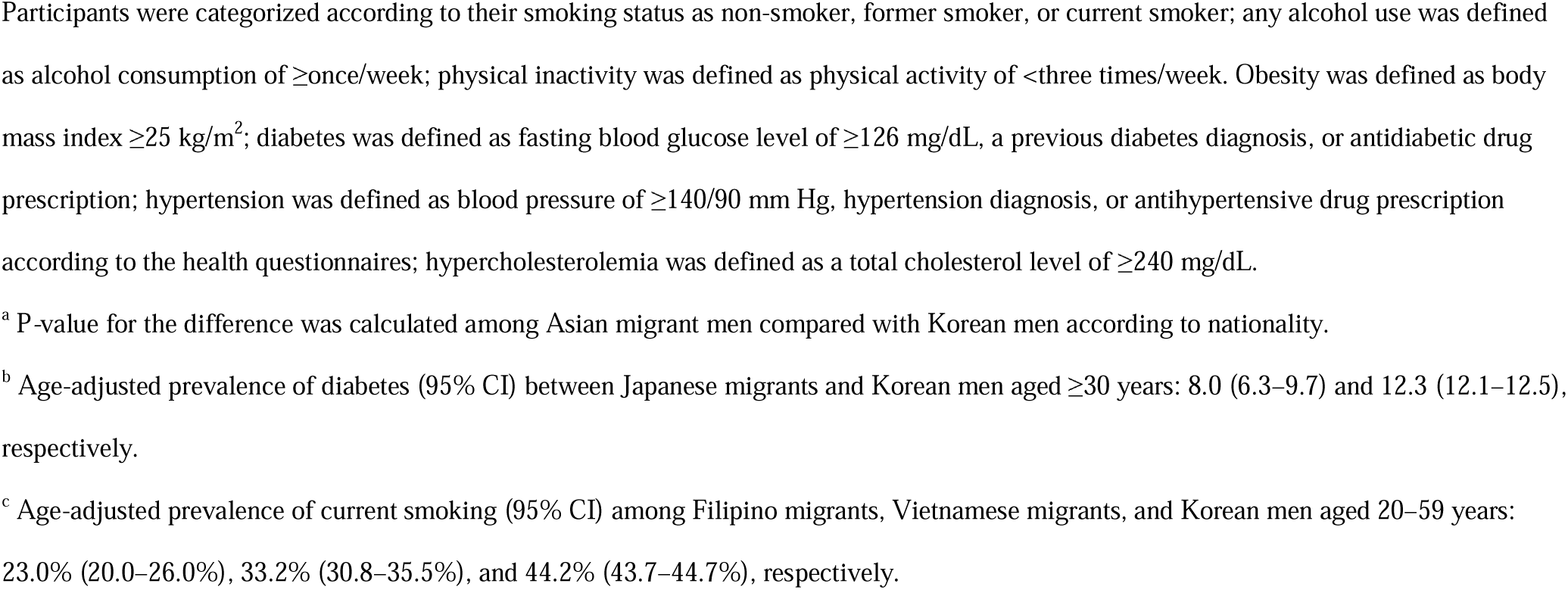
Comparison of health-related indicators between Asian migrant and Korean men ≥20 years in 2015

**Table 3.**
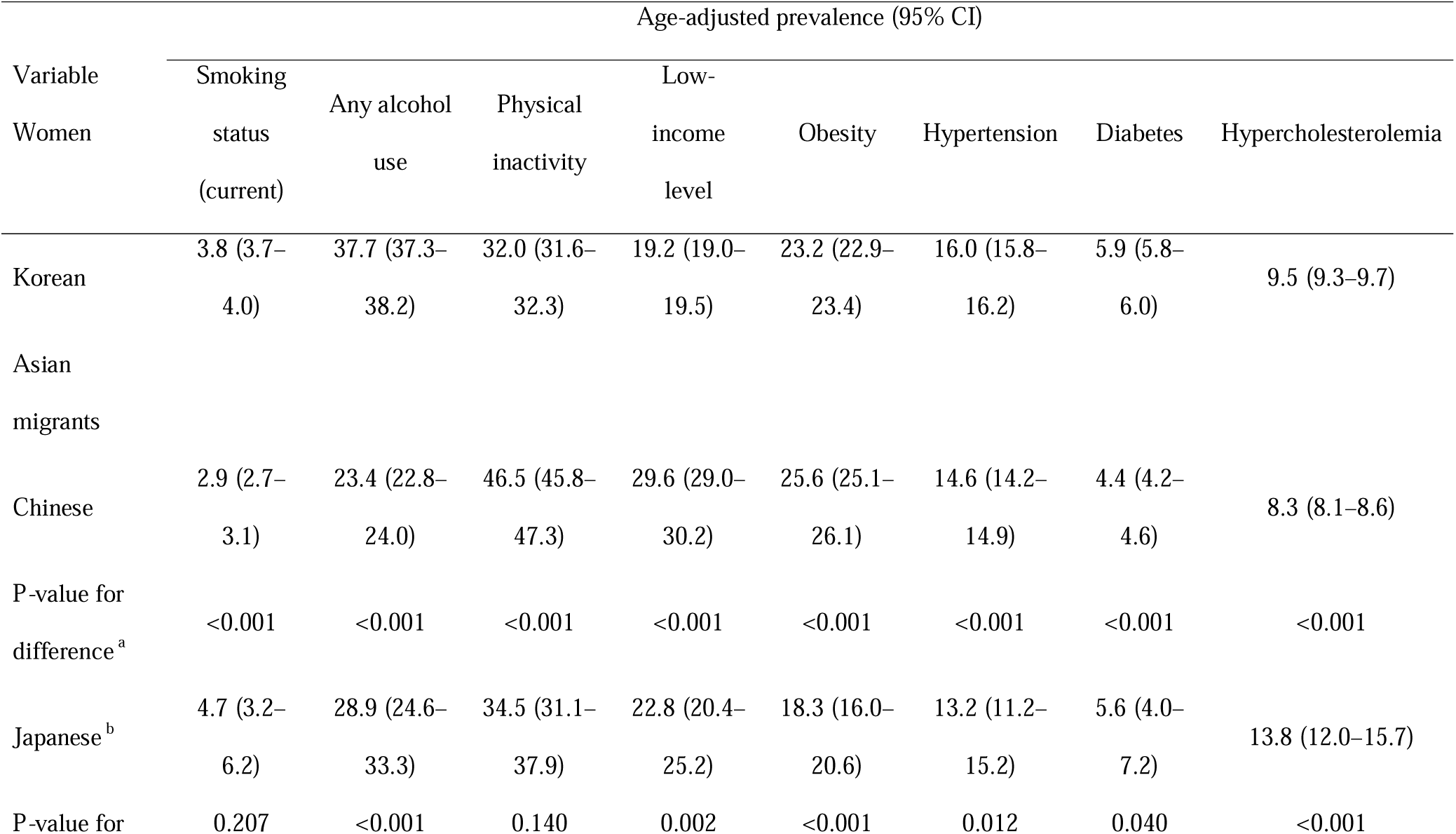
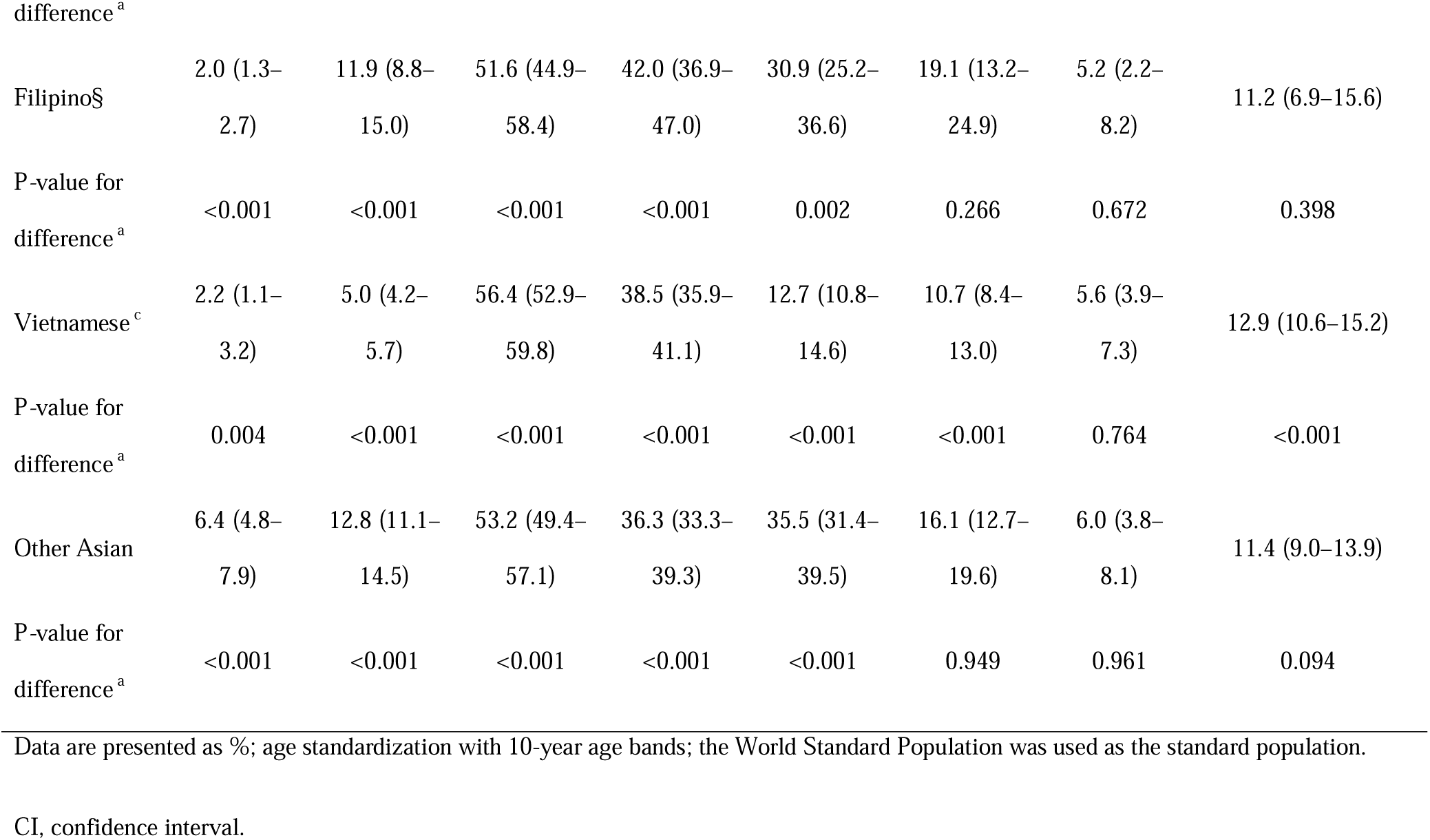
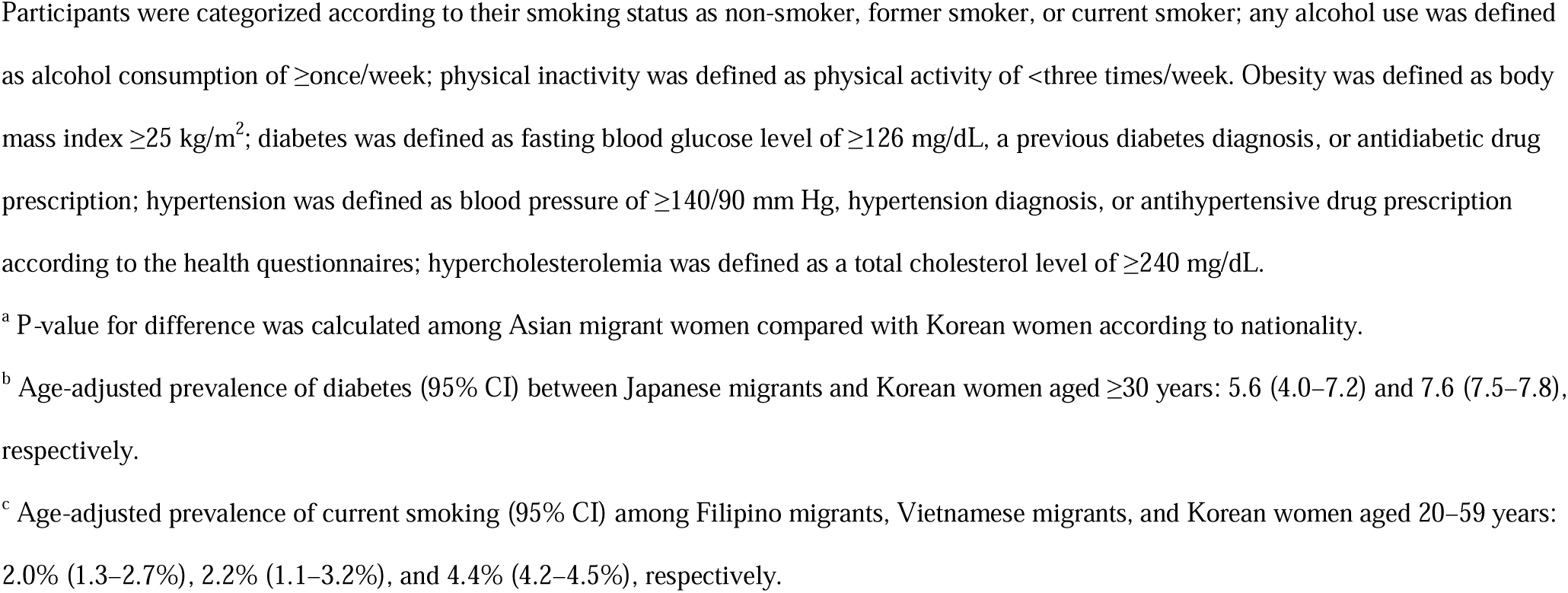
Comparison of health-related indicators between Asian migrant and Korean women aged ≥20 years in 2015

In 2015, among participants aged 30–39 years, the prevalence of diabetes was higher in Chinese migrant men than in Korean men (prevalence ratio [PR], 1.20; 95% CI: 1.07–1.33). Among participants aged 40–49 years, the prevalence of obesity and hypertension was higher in Chinese migrant women than in Korean women (PR, 1.17; 95% CI: 1.13–1.22 and PR, 1.10; 95% CI: 1.04–1.17, respectively) (eTables 1 and 2).

In 2015, among participants aged 20–29 and 30–39 years, the prevalence of hypercholesterolemia was higher in Filipino migrant men than in Korean men (PR, 1.37; 95% CI: 1.18–1.61 and PR, 1.09; 95% CI: 1.00–1.18, respectively). Among participants aged 30– 39 years, the prevalence of diabetes was higher in Filipino migrant women than in Korean women (PR, 2.08; 95% CI: 1.47–2.94) (eTables 3 and 4).

In 2015, among participants aged 20–49 years, the age-adjusted prevalence of hypercholesterolemia was higher in Filipino migrant men than in Korean men (P=0.010), diabetes was higher in Filipino migrant women than in Korean women (aPR, 1.49; 95% CI: 1.11–2.02; P=0.009), and hypertension was higher in Filipino migrant women than in Korean women (aPR, 1.81; 95% CI: 1.48–2.21; P<0.001) (Figure 1 and eTable 5).

**Figure 1.**
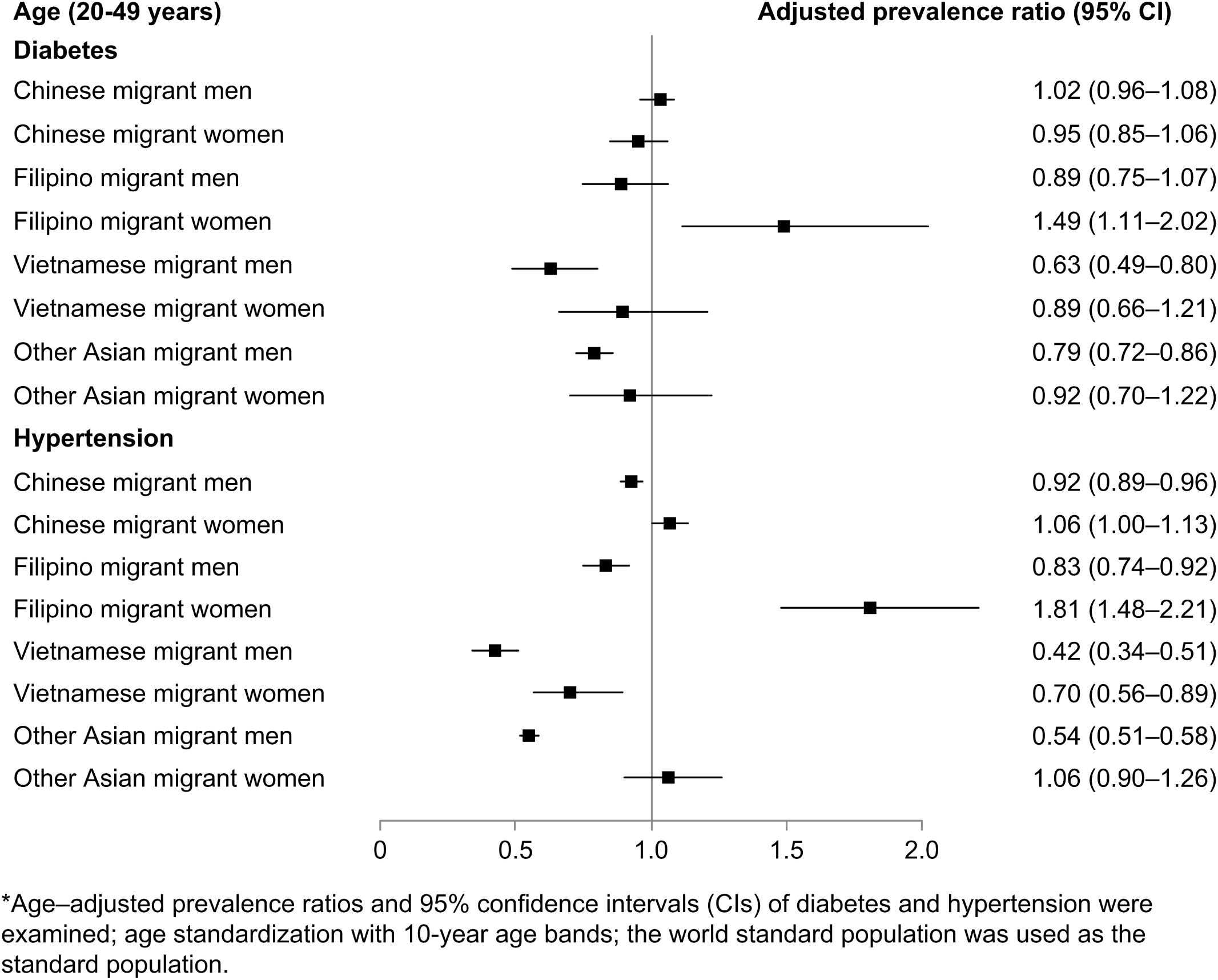
Diabetes and hypertension in Asian migrants versus Koreans aged 20–49 years in 2015*. * Age-adjusted prevalence ratios and 95% confidence intervals (CIs) of diabetes and hypertension were examined, with age standardization using 10-year age bands. The World Standard Population was used as the standard population. Diabetes was defined as fasting blood glucose level of ≥126 mg/dL, a previous diabetes diagnosis, or antidiabetic drug prescription. Hypertension was defined as blood pressure of ≥140/90 mm Hg, hypertension diagnosis, or antihypertensive drug prescription according to the health questionnaires.

During 2009–2015, in multivariable analyses after adjusting for covariates, among participants aged 20–39 years, Vietnamese migrant men were more likely to have T2D than Korean men; among participants aged ≥20 years, Filipino migrant women were more likely to have hypertension than Korean women (eFigures 2 and 3; eResults).

In 2015, the age-adjusted prevalence of obesity was higher in Filipino migrant men than in the general male population in the Philippines. Moreover, in 2014, the age-adjusted prevalence of diabetes was higher in Filipino migrant men than in Filipino men (Figure 2).

**Figure 2.**
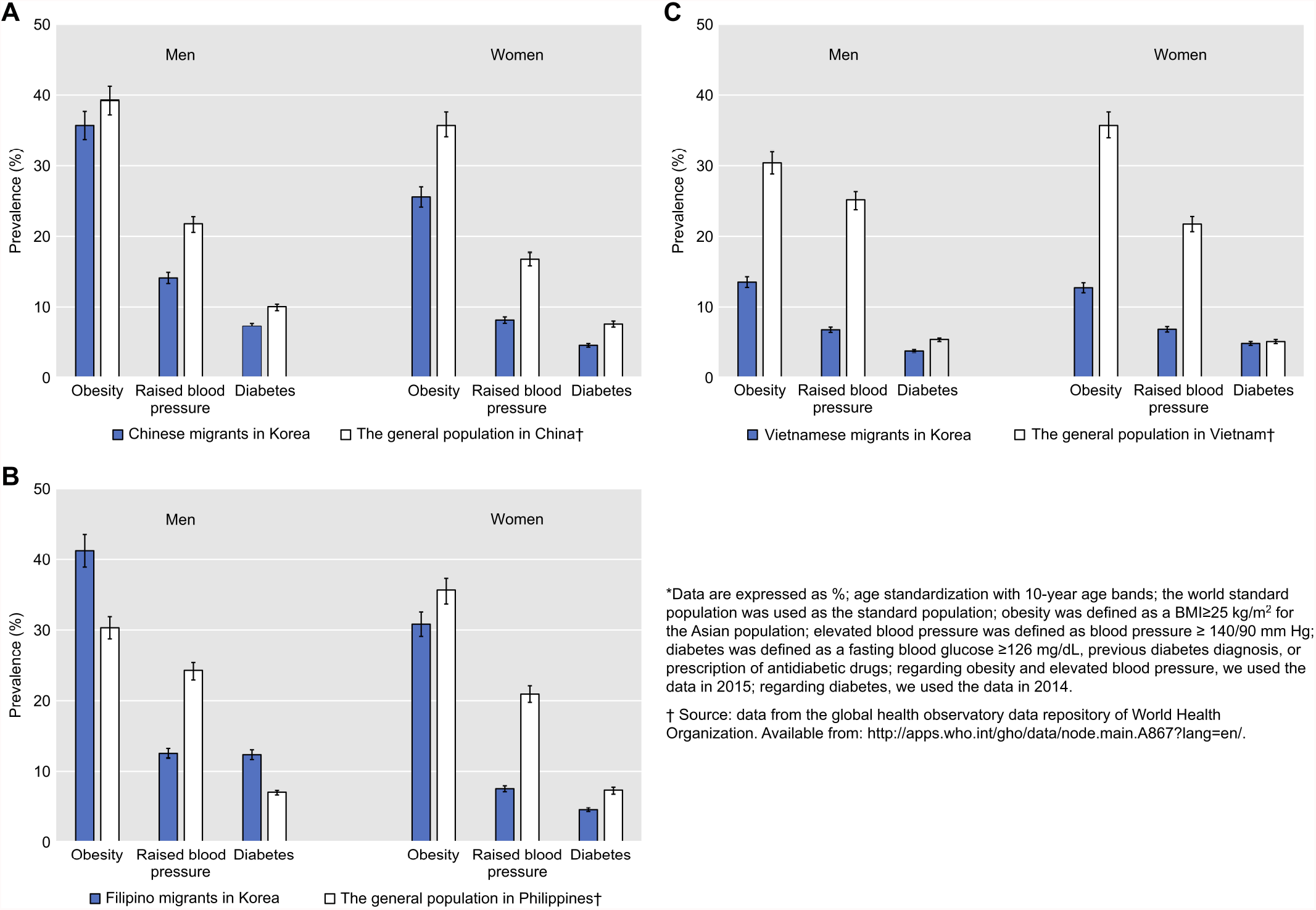
Comparing health-related indicators in Asian migrants and the general population in their home countries*. * Data are presented as %. Age standardization was conducted using 10-year age bands. The World Standard Population was used as the standard population. Obesity was defined as a body mass index ≥25 kg/m^2^ for the Asian population. Elevated blood pressure was defined as blood pressure ≥140/90 mmHg. Diabetes was defined as fasting blood glucose level of ≥126 mg/dL, a previous diabetes diagnosis, or antidiabetic drug prescription. Regarding obesity and elevated blood pressure, 2015 data were used; for diabetes, 2014 data were used.

## Discussion

The age-adjusted prevalence of current smoking was higher in Chinese migrant men than in Korean men and higher in other Asian migrant women than in Korean women aged ≥20 years in 2015. The prevalence of current smoking among migrants is changing according to the situation at home and in the host countries.^19,20^ Smoking is harmful to the health of both the migrant and native populations. Risk factors attributed to social inequality may contribute to more than half of the major NCDs, especially CVD and lung cancer.^14,21,22^

In 2015, the age-adjusted prevalence of obesity was higher in Chinese, Filipino, and other Asian migrant women than in Korean women aged ≥20 years. In our previous study analyzing the characteristics of migrants, the number of women who underwent health check-ups was less than the number of men; however, the number of women increased with each year during 2009–2015.^8^ Obesity is a major health problem, particularly in migrant women.^8,11,23^ Unhealthy lifestyles, socioeconomic and cultural factors, genetics, and gene-environment interactions may lead to obesity, diabetes, and hypertension.^11,24,25^ Compared with Koreans, we could not demonstrate concurrence between obesity and diabetes/hypertension among Asian migrants, consistent with previous findings showing that there was no concurrence between obesity and T2D among international migrants from the Western Pacific to Europe.^23^ Nonetheless, there was concurrence between obesity and hypertension in Chinese migrant women aged 40–49 years. Socioeconomic inequalities, cultural stress, and obesity may be the main factors contributing to hypertension among migrants.^4,14,26^ Among Filipino migrant women aged 20–49 years, we found concurrence between obesity and diabetes/hypertension compared with Korean women, probably because of unhealthy diets, lack of exercise, early life factors, poor adherence to medication, low socioeconomic status, genetics, and gene-environment interactions.^11,24,25^ The stress generated by adapting to a new culture may contribute to a greater prevalence of hypertension among Filipino migrant women aged 20–49 years; this appears to be more important than unhealthy diets or lack of exercise.^26,27^

Here, Japanese and Vietnamese migrants had a higher prevalence of hypercholesterolemia than Koreans. Among participants aged 20–49 years, Filipino migrant men had a higher prevalence of hypercholesterolemia than Korean men, especially those aged 20–29 and 30–39 years. Previous studies have shown that because of Americanized diets, which are higher in fat than traditional Japanese diets, total cholesterol levels became higher in Japanese Americans than in Japanese natives.^28,29^ Michael Marmot’s doctoral thesis on coronary heart disease (CHD) in Japanese Americans illustrated that participants who adhered to Japanese culture and had closer social networks were less likely to develop CHD compared with those who were more accustomed to Western culture and lifestyles.^30^ Although diet and hypercholesterolemia may increase the prevalence of CHD, the protective effect of Japanese culture on CHD is important, irreplaceable, and even independent of unhealthy diets and smoking.^28,30,31^ Therefore, the protective effect of social and cultural factors on cardiovascular health should be addressed while advocating healthy lifestyles.

In 2015, Chinese migrant men had a greater prevalence of diabetes than Korean men, whereas Filipino migrant women had a greater prevalence of diabetes than Korean women aged 30–39 years. Migrants aged 30–39 years compared with Koreans may be more susceptible to diabetes after several migrant-related lifestyle changes.^7,11,25^ However, in multivariable analyses, during 2009–2015 among participants aged 20–39 years, Vietnamese migrant men were more likely to have T2D than Korean men. This finding is consistent with previous studies showing that international migrants developed diabetes approximately 10–20 years earlier than the local population of their host countries.^7,11,24^ Vietnamese migrant men aged 20–39 years may be more susceptible to T2D because of greater vulnerability to stress from Korean society and culture, under the combined effect of unhealthy diets, genetics, and gene-environment interactions.^14,32^

Considering the important relationship between the pressure of cultural changes and elevated blood pressure among migrants, the reason why most Asian migrants are less likely to develop hypertension than Koreans may be due to the similar cultures and relatively close proximity to their home country.^11,26,27^ Asian migrants may be more likely to adapt to Korean society and maintain large social networks, ensuring sufficient social support. These factors may help Asian migrants in Korea reduce stress and deal with conflicts more easily than when adapting to a different culture. Nevertheless, we cannot exclude the possibility that Asian migrants may return to their home countries either because of poor health or financial difficulties in Korea.^19,33^ Therefore, while the overall incidence of diabetes and hypertension was lower among Asian migrants in this and previous studies, the true incidence may be higher. However, Filipino migrant women were more likely to develop hypertension than Korean women, possibly due to social and cultural challenges encountered by Filipino women during the process of acculturation to Korea.^26,27^

Compared with the general population in their home countries, most Asian migrants in Korea were healthier regarding the prevalence of obesity, diabetes, and elevated blood pressure.^11^ Nevertheless, the prevalence of obesity and diabetes was higher among Filipino migrant men than among the general male Philippine population, potentially due to unhealthy lifestyles, low socioeconomic status, genetics, and gene-environment interactions.^11,24,25^ During 2009– 2015, the age-adjusted prevalence of diabetes, hypertension, and hypercholesterolemia was lower among most Asian migrants—regardless of sex—than among Koreans.^8^ Based on previous studies, the pursuit for better economic, educational, or social and living environment needs, combined with the self-selection of migrants and the immigration policy of the Korean government, may explain the healthy migration advantage in some NCD risk factors among most Asian migrants in Korea.^1,2,34^ However, regarding Israel, the reason for the sick migration effect may be attributed to the Jewish population’s international migration to Israel for ideological and not economic motivations compared with the healthy migration effect in European countries.^5^

Our study has several strengths. This was a large-scale observational study comparing the prevalence of NCD risk factors among Asian migrants of different nationalities and the host population in Korea. The design was cross-sectional in 2015 and longitudinal during 2009– 2015 (eDiscussion). Few similar studies have investigated the healthy migration effect among international migrant populations of various ethnic groups within the host populations of high-income countries in Asia using large data from the same source. Because of the lack of complete standardized worldwide migrant data, even a global-scale study that verified the healthy migration effects in high-income countries could only be conducted by pooling mortality data and considering heterogeneity (high-quality data from different sources or sectors) and did not include international migrants in Asia.^2^ Therefore, the innovative point here is that, by comparing Asian migrants with Korean natives using health check-up data from the Korean NHIS, we conducted age standardization using the World Standard Population and compared the health data of Asian migrants with the health data in their home countries, targeting the general populations of China, the Philippines, and Vietnam, published by the Global Health Observatory Data Repository of the World Health Organization.^17^

Our study has several limitations. First, the total number of diabetes and hypertension cases was underestimated because we only selected individuals who had National Health Insurance and underwent health check-ups. Further, the study population could have been healthier than the entire population if the individuals were more attentive to their health and fully used medical resources. However, a previous study using the same data (during 2009–2015) showed that the prevalence of diabetes in Korean men and women aged ≥20 years showed an increasing trend, in line with the results of a recent study on Korean adults aged ≥30 years based on the Korean National Health and Nutrition Examination Survey.^8,35^ Moreover, the prevalence of diabetes and hypertension is more likely to have been underestimated in Asian migrants than in Koreans.^11^ For example, in Singapore, although international migrant workers have National Health Insurance when employed, they may not utilize necessary health services or even know that they have been covered.^36^ Second, we did not adjust for family history, nutritional factors, stress, and depression in the statistical analysis of relationships among Asian migrants according to nationality and newly diagnosed T2D or hypertension compared with Koreans. Third, because of the limited use of data on the type of international migrants (migrant workers, international students, and marriage migrants), migrant status factors were not considered; however, we used economic status instead of migrant status to compensate^8^ (eDiscussion).

In conclusion, considering the adverse effects of socioeconomic and cultural factors on public health, strategies are needed to target smoking and obesity among Asian migrants of specific nationalities in Korea to address primary and secondary prevention of NCD, especially CVD and lung cancer. Moreover, health inequalities exist among Filipino migrants, especially women aged 20–49 years.

## Data Availability

No additional data were available.

https://nhiss.nhis.or.kr/bd/ab/bdaba000eng.do;jsessionid=xIoMvELC4gUI0wBh01wHeTUmP1oRqukCUszPBvoRy1JptkJeVJHKa3wGp5s9Jo7h.primrose22_servlet_engine1

## Author Contributions

HP, JMY, AS, and BC conceived the study. HP wrote the first and successive drafts of the manuscript. HP modeled and analyzed the data. HP, JMY, and AS contributed to study conception and design. JMY, AS, and DK contributed to data analysis. All authors revised the manuscript for important intellectual content. HP and BC had full access to the data and take responsibility for the integrity of the data and the accuracy of the data analysis. HP and BC are the guarantors. The corresponding author attests that all the listed authors meet the authorship criteria and that no others meeting the criteria have been omitted.

## Conflicts of Interest Disclosures

Belong Cho reports grants and non-financial support from Yuhan Pharmaceutical during the conduct of the study. Heng Piao reports grants and non-financial support from Yuhan Pharmaceutical during the conduct of the study. The remaining authors have no financial relationships with any organizations that might have an interest in the submitted work in the previous 3 years; there are no other relationships or activities that could influence the submitted work. The authors had access to all the study data, take responsibility for the accuracy of the analysis, and had authority over manuscript preparation and the decision to submit the manuscript for publication. All authors approve the manuscript and agree to adhere to all terms outlined in Annals of Internal Medicine Information for Authors, including terms for copyright.

## Funding Source

This study was funded by Seoul National University Hospital, Seoul, Republic of Korea; Yuhan Pharmaceutical, Seoul, Republic of Korea; and Henan Cancer Hospital/Affiliated Cancer Hospital of Zhengzhou University, Zhengzhou, China.

## Meeting Presentation

Accepted Oral Presentation at the 23rd World Conference of Family Doctors 2021 (WONCA 2021). WONCA 2021 was from November 22, 2021, to November 27, 2021.

## Role of the Funding Source

The funders had no role in the design of the study; the collection, analysis, and interpretation of the data; or the decision to approve publication of the finished manuscript.

## Additional Contributions

We thank Carol Brayne and Caroline Lee (University of Cambridge), Jae-Heon Kang (Sungkyunkwan University), Jian Li (University of California), Jong-Koo Lee and Young-Ho Khang (Seoul National University), and Shu-Zheng Liu (Zhengzhou University) for their constructive suggestions and comments. We also thank Editage (www.editage.cn), Luyao Zhang (Zhengzhou University), and Mohamed S. Bangura (Dalian Medical University) for their writing support.

## Notes

### Competing Interest Statement

All authors have completed the ICMJE uniform disclosure form at www.icmje.org/coi_disclosure.pdf and declare that: Belong Cho reports grants and non-financial support from Yuhan Pharmaceutical, during the conduct of the study. Heng Piao reports grants and non-financial support from Yuhan Pharmaceutical, during the conduct of the study. The other authors have no financial relationships with any organizations that might have an interest in the submitted work in the previous 3 years; and that there are no other relationships or activities that could appear to have influenced the submitted work.

### Clinical Trial

NHIS-2021-1-008

### Funding Statement

This study was funded by the Seoul National University Hospital (Republic of Korea), Yuhan Pharmaceutical (Republic of Korea), and Henan Cancer Hospital/Affiliated Cancer Hospital of Zhengzhou University (China). The funders had no role in study design, data collection and analysis, decision to publish, or preparation of the manuscript.

### Author Declarations

The Institutional Review Board of Seoul National University Hospital approved the study on August 21, 2018 (IRB number: E-1808-093-966). Moreover, per the criteria of IRB, the need for obtaining informed participant consent was waived in our study.

### Summary of Updates

Section on Manuscript updated to clarify Title, Abstract, Introduction, Methods, Results, and Discussion; Section on Manuscript updated to clarify Conclusion; Supplemental files updated; etc.

