## Supplementary Appendix for "Comparing Non-Communicable Disease Risk Factors in Asian Migrants and Native Koreans"

**Online-Only Supplement**

**List of Online-Only Supplement**

- **Online-Only Text**

**eMethods**

**eResults**

**eDiscussion**

- **Online-Only Figures**

**eFigure 1.** Flow diagram of the research design and statistical analysis

**eFigure 2.** Type 2 diabetes development comparing Asian migrants of different nationalities with Koreans during 2009–2015*

**eFigure 3.** Hypertension development comparing Asian migrants of different nationalities with Koreans during 2009–2015*

- **Online-Only Tables**

**eTable 1.** Health-related indicators comparing Chinese migrant men with Korean men aged 20–49 years in 2015

**eTable 2.** Health-related indicators comparing Chinese migrant women with Korean women aged 20–49 years in 2015

**eTable 3.** Health-related indicators comparing Filipino migrant men with Korean men aged 20–49 years in 2015

**eTable 4.** Health-related indicators comparing Filipino migrant women with Korean women aged 20–49 years in 2015

**eTable 5.** Comparing age-adjusted health-related indicators between Filipino migrants and Koreans aged 20–49 years in 2015

- **Online-Only References**

**Online-Only Text**

**eMethods**

Among 2,691,010 participants aged ≥20 years who underwent health check-ups between 2009 and 2015, we excluded 350 lacking information on blood pressure, fasting blood glucose, and total cholesterol levels. Furthermore, 10,165 individuals with missing information on smoking status, alcohol use, or physical activity were excluded. Finally, the study population of 2,680,495 participants included 987,214 Asian migrants and 1,693,281 Republic of Korea nationals.[1]

Between 2009 and 2015, the numbers of Asian migrants by year were 93,845, 113,414, 129,827, 132,226, 142,275, 175,082, and 200,545, respectively. Similarly, the numbers of Koreans by year were 208,772, 231,881, 236,751, 244,443, 243,592, 261,680, and 266,162, respectively.[1]

In 2015, the mean (standard deviation) age among Korean natives and Chinese, Japanese, Filipino, Vietnamese, and other Asian migrants was 49.4 (14.0), 46.4 (11.4), 47.4 (9.2), 32.5 (6.0), 30.3 (9.2), and 30.3 (6.3) years, respectively. In addition, the number of women among Korean natives, Chinese, Japanese, Filipino, Vietnamese, and other Asian migrants was 124,850 (46.9%), 47,808 (47.2%), 3,171 (71.3%), 2,347 (19.7%), 5,600 (29.1%), and 4,648 (7.3%) years, respectively (Table 1).

For the accuracy of age standardization, migrants from Thailand, Uzbekistan, Sri Lanka, Mongolia, Bangladesh, Pakistan, Indonesia, and India were labeled “other Asian migrants” when the data were derived from the National Health Insurance Service (NHIS). For example, there were insufficient samples for several positive indicators of non-communicable disease (NCD) risk factors among Indonesian migrants aged ≥60 years. Therefore, migrants from Thailand, Uzbekistan, Sri Lanka, Mongolia, Bangladesh, Pakistan, Indonesia, and India were combined as “other Asian migrants” collectively, as age standardization resulted in insufficient data on specific positive outcome indicators.[2]

According to nationality, there were insufficient samples among Asian migrants aged 60–69, 70–79, and >80 years for age standardization by sex for several positive indicators of

NCD risk factors, especially among Filipino, Vietnamese, and other Asian migrants. Therefore, the study population was divided into age groups of 20–29, 30–39, 40–49, 50–59, and >60 years to conduct age standardization for NCD risk factors among Asian migrants versus Koreans according to nationality.

The health check-up participation data were obtained from the NHIS Health Check-Up Database. We used monthly insurance premiums to indicate economic status (first, second, third, and fourth quartiles), which was regarded as a vital statistic that represented income level (low, middle-low, high-middle, and high, respectively). Based on the information obtained from the health questionnaires, participants were categorized according to their smoking status as non-smoker, former smoker, or current smoker; any alcohol use was defined as alcohol consumption of ≥once/week; participants were categorized to be physically active if physical activity was ≥three times/week or physically inactive if physical activity was <three times/week. Weight, height, and blood pressure measurements were included in the physical examinations, and body mass index (BMI) was calculated as the weight (kg) divided by the height ($m^2$). Obesity was defined as a BMI of ≥25 kg/$m^2$. Elevated blood pressure (hypertension) was defined as blood pressure ≥140/90 mm Hg, a diagnosis of hypertension, or prescription of antihypertensive drugs in the health questionnaires. Furthermore, total cholesterol and fasting blood glucose were included in the laboratory tests. Hypercholesterolemia was defined as a total cholesterol level of ≥240 mg/dL. Diabetes was defined as a fasting blood glucose level of ≥126 mg/dL, a previous diabetes diagnosis, or prescription of antidiabetic drugs in the health questionnaires.[2]

The age-adjusted prevalence of lifestyle, socioeconomic, and health-related factors among Asian migrants and the native Korean population, according to nationality, was calculated. The age-adjusted prevalence was calculated separately by sex using direct age standardization. However, to compare the age-adjusted prevalence of diabetes among Japanese migrants with that of Koreans, we selected Japanese migrant and Korean populations aged ≥30 years, as there were insufficient samples among Japanese migrants aged 20–29 years for age standardization in diabetes. Moreover, we selected the Filipino and Vietnamese populations and compared them with the Korean population aged 20–59 years for age standardization of smoking because there were insufficient participants in the category of Filipino and Vietnamese migrants aged ≥60 years. Furthermore, considering the limitation of overall insufficient samples among Asian migrants of different nationalities aged 50–59, 60–69, 70–79, and >80 years, we calculated the age-specific and age-adjusted prevalence of lifestyle, socioeconomic, and health-related factors among Chinese, Filipino, Vietnamese, and other Asian migrants compared with Koreans, separately by sex, using 10-year age bands for the ages 20–49 years. Regarding direct age standardization for the ages 20–49 years, Japanese migrants aged 20–29 years were excluded owing to insufficient data. In addition, for the health of Asian migrants compared with the general population in their home countries, Japanese migrants were excluded because of insufficient data of Japanese migrants aged 20–29 years.

**eResults**

During 2009–2015, the number of participants who underwent health check-ups more than once with available data regarding diabetes was 505,342. The number of participants with incident type 2 diabetes (T2D) between the first and last health check-ups was 22,284. In multivariate analyses, compared with native Koreans, the odds ratio (OR) for developing T2D after adjusting for covariates was 0.82 (95% confidence interval [CI]: 0.78–0.86) among

Asian migrants. However, among Vietnamese migrant men aged 20–39 years, the OR for developing T2D was 1.32 (95% CI: 1.11–1.57) compared with Korean men (eFigure 2).

During 2009–2015, the number of participants who underwent health check-ups more than once with available data regarding hypertension was 431,433. The number of participants with hypertension between the first and last health check-ups was 48,007. In multivariate analyses, compared with native Koreans, the OR for developing hypertension after adjusting for covariates was 0.77 (95% CI: 0.75–0.79) among Asian migrants. However, compared with Korean women, the ORs for developing hypertension were 1.49 (95% CI: 1.05–2.11) and 2.22 (95% CI: 1.17–4.19) among Filipino migrant women aged 20–39 and >40 years, respectively (eFigure 3).

**eDiscussion**

In our study, the design was cross-sectional in 2015 and longitudinal during 2009–2015. Longitudinal data were collected to examine the differences in cases of newly diagnosed T2D and hypertension between Asian migrants according to nationality and Koreans, after adjusting for lifestyle factors. Regarding the longitudinal analysis, a recent review has addressed the importance of answering research questions on migrant health using longitudinal data.[3]

We could not distinguish between essential and secondary hypertensive participants, although secondary hypertension is only expected in 5–10% of participants with hypertension, which may not affect our conclusions.[4,5] In addition, there were insufficient samples among Japanese, Filipino, and Vietnamese migrants regarding several NCD risk factors of a specific age group. Nevertheless, we overcame these limitations by using cross-sectional and longitudinal designs based on age, sex, and socioeconomic status of Asian migrants of different nationalities in Korea. Finally, we were unaware of their residence duration and could not calculate the exact follow-up time of incident T2D and hypertension. However, even using the Kaplan-Meier method with adequate follow-up time, it was impossible to accurately compare changes in diabetes and hypertension over time in multiple populations of different nationalities during 2009–2015. This results from differences in countries of birth, early and lifestyle factors, and cultural backgrounds between Asian migrants and Korean natives. However, we performed an annual trend analysis of lifestyle, socioeconomic, and health-related factors among Asian migrants and Koreans during 2009–2015 in our previous study.[1] These trend analyses provide insights into changes in the overall prevalence of obesity, diabetes, and hypertension among Asian migrants compared with Koreans over time.

### Online-Only Figures

### eFigure 1. Flow diagram of the research design and statistical analysis

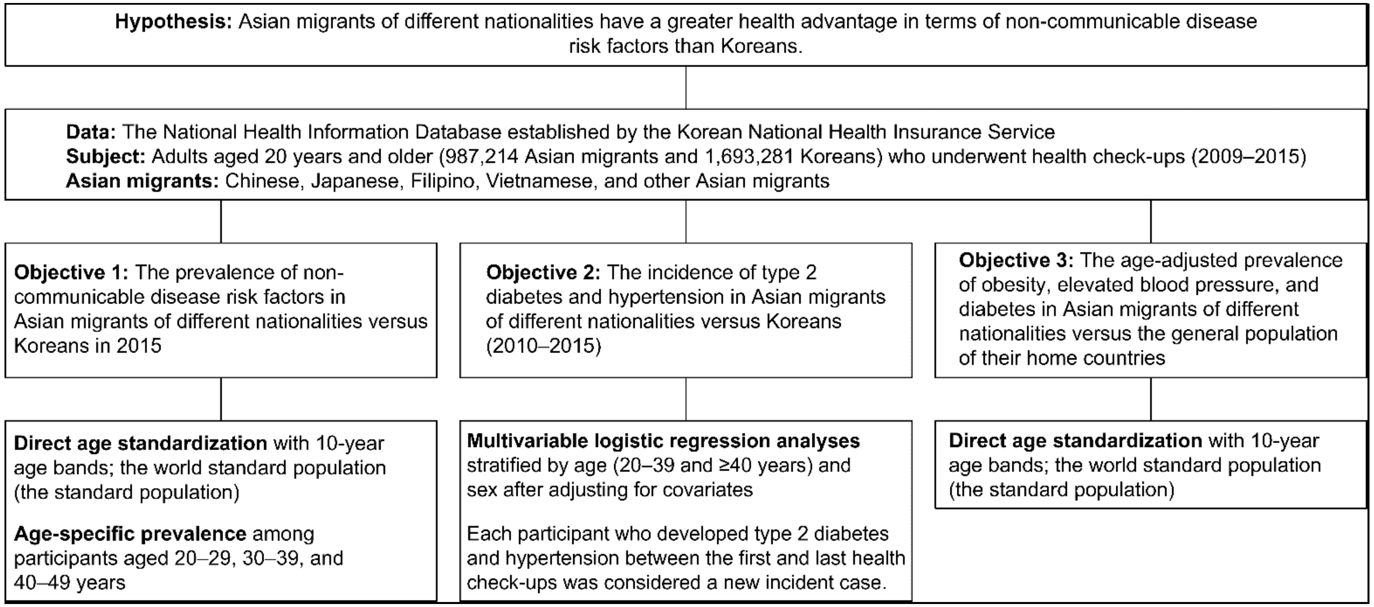

**eFigure 2. Type 2 diabetes development comparing Asian migrants of different nationalities with Koreans during 2009–2015***

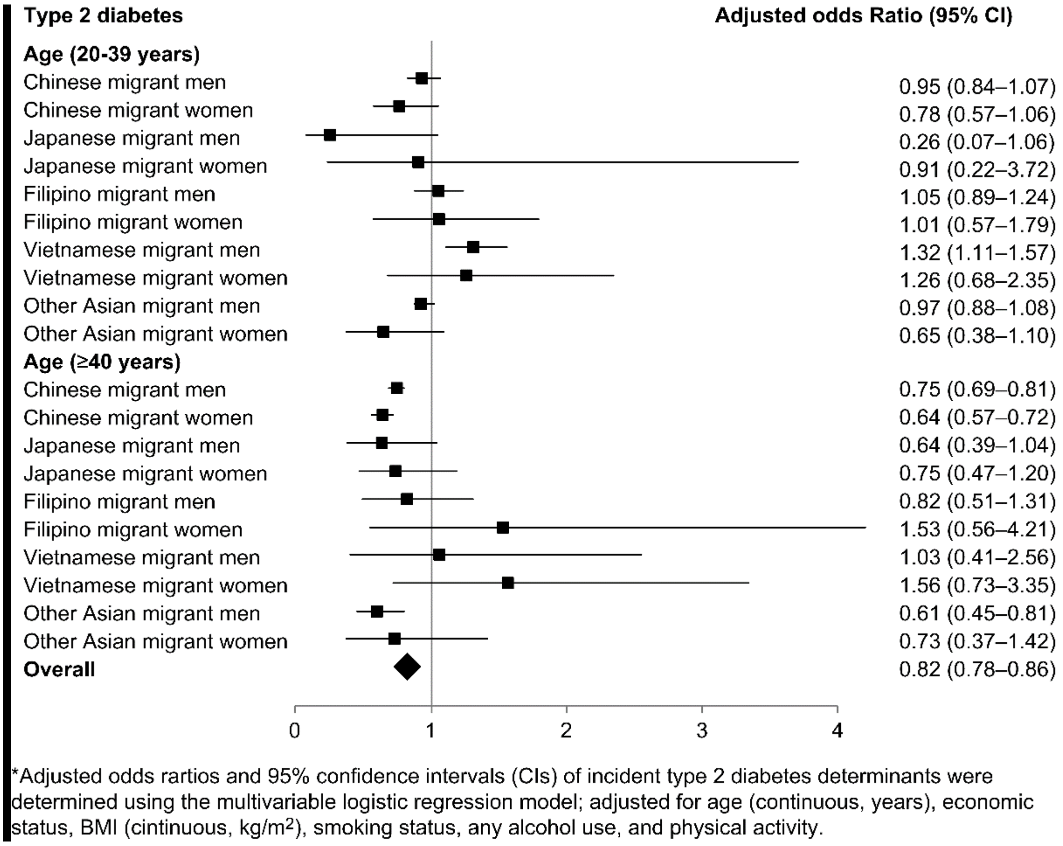


* Adjusted odds ratios and 95% confidence intervals (CIs) of incident type 2 diabetes determinants were determined using the multivariable logistic regression model that was adjusted for age (continuous, years), economic status, body mass index (continuous, kg/$m^2$), smoking status, any alcohol use, and physical activity.

**eFigure 3. Hypertension development comparing Asian migrants of different nationalities with Koreans during 2009–2015***

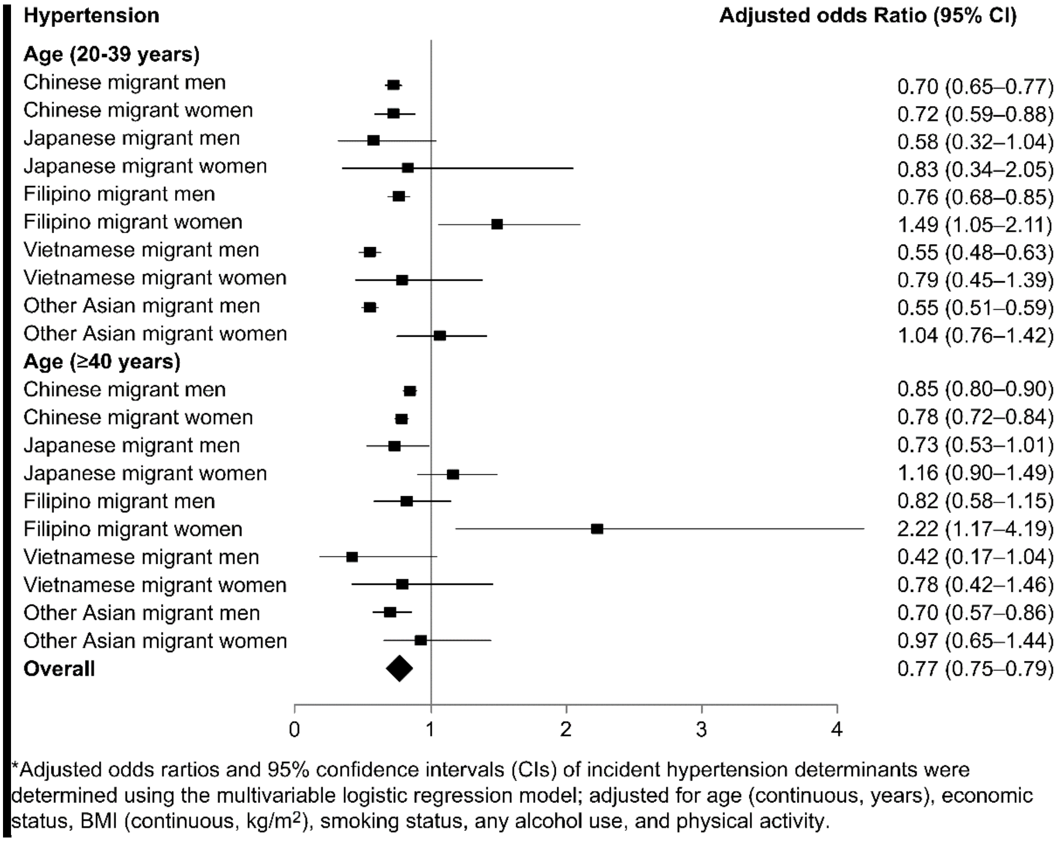


* Adjusted odds ratios and 95% confidence intervals (CIs) of incident hypertension determinants were determined using the multivariable logistic regression model that was adjusted for age (continuous, years), economic status, body mass index (continuous, $kg/m^2$), smoking status, any alcohol use, and physical activity.

**Online-Only Tables**

**eTable 1.** Health-related indicators comparing Chinese migrant men with Korean men aged 20–49 years in 2015

| **Variable** | **Age-adjusted prevalence**[a] | | **Prevalence ratio** | **95% lognormal confidence interval** | |
|---|---|---|---|---|---|
| | **Chinese migrant men** | **Korean men** | **Chinese migrant men *versus* Korean men** | **Lower limit** | **Upper limit** |
| Smoking status (current) | | | | | |
| 20–29 years | 60.7 | 46.5 | 1.30 | 1.25 | 1.36 |
| 30–39 years | 61.6 | 47.6 | 1.29 | 1.26 | 1.33 |
| 40–49 years | 54.2 | 45.1 | 1.20 | 1.17 | 1.23 |
| Any alcohol use | | | | | |
| 20–29 years | 65.9 | 77.0 | 0.86 | 0.82 | 0.89 |
| 30–39 years | 72.6 | 76.6 | 0.95 | 0.92 | 0.97 |

| | | | | | |
|---|---|---|---|---|---|
| 40–49 years | 74.0 | 74.1 | 1.00 | 0.98 | 1.02 |
| Physical inactivity | | | | | |
| 20–29 years | 42.6 | 18.7 | 2.28 | 2.14 | 2.42 |
| 30–39 years | 44.7 | 24.0 | 1.86 | 1.79 | 1.93 |
| 40–49 years | 50.7 | 27.3 | 1.86 | 1.80 | 1.91 |
| Low income level | | | | | |
| 20–29 years | 23.5 | 13.0 | 1.81 | 1.68 | 1.95 |
| 30–39 years | 15.3 | 4.5 | 3.38 | 3.15 | 3.63 |
| 40–49 years | 16.5 | 8.5 | 1.95 | 1.85 | 2.05 |
| Obesity | | | | | |
| 20–29 years | 31.9 | 37.5 | 0.85 | 0.80 | 0.90 |
| 30–39 years | 38.4 | 46.9 | 0.82 | 0.79 | 0.85 |
| 40–49 years | 38.2 | 45.8 | 0.83 | 0.81 | 0.86 |
| Hypertension | | | | | |
| 20–29 years | 5.0 | 6.1 | 0.82 | 0.71 | 0.94 |

| | | | | | |
|---|---|---|---|---|---|
| 30–39 years | 9.6 | 11.2 | 0.86 | 0.80 | 0.92 |
| 40–49 years | 21.0 | 21.1 | 1.00 | 0.96 | 1.04 |
| Diabetes | | | | | |
| 20–29 years | 1.6 | 1.4 | 1.11 | 0.85 | 1.44 |
| 30–39 years | 4.1 | 3.5 | 1.20 | 1.07 | 1.33 |
| 40–49 years | 8.3 | 9.0 | 0.92 | 0.86 | 0.98 |
| Hypercholesterolemia | | | | | |
| 20–29 years | 4.4 | 5.4 | 0.82 | 0.71 | 0.95 |
| 30–39 years | 7.4 | 11.8 | 0.63 | 0.59 | 0.68 |
| 40–49 years | 9.4 | 14.9 | 0.63 | 0.60 | 0.67 |

[a] Data are expressed as percentages.

**eTable 2.** Health-related indicators comparing Chinese migrant women with Korean women aged 20–49 years in 2015

| Variable | Age-adjusted prevalence [a] | | Prevalence ratio | 95% lognormal confidence interval | |
|---|---|---|---|---|---|
| | Chinese migrant women | Korean women | Chinese migrant women *versus* Korean women | Lower limit | Upper limit |
| Smoking status (current) | | | | | |
| 20–29 years | 4.7 | 6.3 | 0.75 | 0.64 | 0.88 |
| 30–39 years | 4.4 | 4.1 | 1.08 | 0.94 | 1.24 |
| 40–49 years | 2.3 | 3.6 | 0.65 | 0.58 | 0.73 |
| Any alcohol use | | | | | |
| 20–29 years | 37.1 | 59.7 | 0.62 | 0.59 | 0.66 |
| 30–39 years | 32.3 | 47.0 | 0.69 | 0.66 | 0.72 |
| 40–49 years | 22.0 | 37.6 | 0.59 | 0.56 | 0.61 |
| Physical inactivity | | | | | |
| 20–29 years | 40.3 | 25.4 | 1.58 | 1.49 | 1.68 |

| | | | | | |
|---|---|---|---|---|---|
| 30–39 years | 47.1 | 33.4 | 1.41 | 1.35 | 1.47 |
| 40–49 years | 51.7 | 32.6 | 1.58 | 1.54 | 1.63 |
| Low income level | | | | | |
| 20–29 years | 34.1 | 16.7 | 2.04 | 1.90 | 2.18 |
| 30–39 years | 28.0 | 15.6 | 1.80 | 1.70 | 1.91 |
| 40–49 years | 28.6 | 22.1 | 1.29 | 1.24 | 1.34 |
| Obesity | | | | | |
| 20–29 years | 12.8 | 12.0 | 1.07 | 0.97 | 1.18 |
| 30–39 years | 22.3 | 16.6 | 1.35 | 1.26 | 1.43 |
| 40–49 years | 27.5 | 23.5 | 1.17 | 1.13 | 1.22 |
| Hypertension | | | | | |
| 20–29 years | 1.2 | 1.3 | 0.95 | 0.69 | 1.31 |
| 30–39 years | 2.8 | 2.8 | 1.00 | 0.84 | 1.19 |
| 40–49 years | 10.7 | 9.7 | 1.10 | 1.04 | 1.17 |
| Diabetes | | | | | |

| | | | | | |
|---|---|---|---|---|---|
| 20–29 years | 0.6 | 0.7 | 0.77 | 0.49 | 1.23 |
| 30–39 years | 1.6 | 1.5 | 1.04 | 0.83 | 1.31 |
| 40–49 years | 3.2 | 3.4 | 0.94 | 0.85 | 1.05 |
| Hypercholesterolemia | | | | | |
| 20–29 years | 2.0 | 3.1 | 0.65 | 0.52 | 0.83 |
| 30–39 years | 3.7 | 6.0 | 0.63 | 0.55 | 0.72 |
| 40–49 years | 8.0 | 8.8 | 0.90 | 0.84 | 0.97 |

[a] Data are expressed as percentages.

**eTable 3.** Health-related indicators comparing Filipino migrant men with Korean men aged 20–49 years in 2015

| Variable | Age-adjusted prevalence [a] | | Prevalence ratio | 95% lognormal confidence interval | |
|---|---|---|---|---|---|
| | Filipino migrant men | Korean men | Filipino migrant men *versus* Korean men | Lower limit | Upper limit |
| Smoking status (current) | | | | | |
| 20–29 years | 27.7 | 46.5 | 0.60 | 0.55 | 0.64 |
| 30–39 years | 23.0 | 47.6 | 0.48 | 0.46 | 0.51 |
| 40–49 years | 18.0 | 45.1 | 0.40 | 0.33 | 0.48 |
| Any alcohol use | | | | | |
| 20–29 years | 55.0 | 77.0 | 0.71 | 0.68 | 0.75 |
| 30–39 years | 54.5 | 76.6 | 0.71 | 0.68 | 0.74 |
| 40–49 years | 50.5 | 74.1 | 0.68 | 0.61 | 0.76 |
| Physical inactivity | | | | | |
| 20–29 years | 51.3 | 18.7 | 2.70 | 2.56 | 2.94 |

| | | | | | |
|---|---|---|---|---|---|
| 30–39 years | 48.7 | 24.0 | 2.04 | 1.96 | 2.13 |
| 40–49 years | 44.0 | 27.3 | 1.61 | 1.43 | 1.82 |
| Low income level | | | | | |
| 20–29 years | 41.9 | 13.0 | 3.23 | 3.03 | 3.45 |
| 30–39 years | 29.0 | 4.5 | 6.25 | 5.88 | 6.67 |
| 40–49 years | 14.3 | 8.5 | 1.69 | 1.35 | 2.08 |
| Obesity | | | | | |
| 20–29 years | 29.0 | 37.5 | 0.78 | 0.72 | 0.83 |
| 30–39 years | 43.1 | 46.9 | 0.92 | 0.88 | 0.96 |
| 40–49 years | 50.5 | 45.8 | 1.10 | 0.98 | 1.23 |
| Hypertension | | | | | |
| 20–29 years | 5.8 | 6.1 | 0.95 | 0.80 | 1.12 |
| 30–39 years | 8.8 | 11.2 | 0.79 | 0.72 | 0.87 |
| 40–49 years | 16.9 | 21.1 | 0.80 | 0.65 | 0.98 |
| Diabetes | | | | | |

| | | | | | |
|---|---|---|---|---|---|
| 20–29 years | 1.3 | 1.4 | 0.93 | 0.65 | 1.32 |
| 30–39 years | 3.9 | 3.5 | 1.14 | 0.98 | 1.30 |
| 40–49 years | 7.0 | 9.0 | 0.78 | 0.57 | 1.06 |
| Hypercholesterolemia | | | | | |
| 20–29 years | 7.4 | 5.4 | 1.37 | 1.18 | 1.61 |
| 30–39 years | 12.7 | 11.8 | 1.09 | 1.00 | 1.18 |
| 40–49 years | 15.8 | 14.9 | 1.06 | 0.86 | 1.30 |

[a] Data are expressed as percentages.

**eTable 4.** Health-related indicators comparing Filipino migrant women with Korean women aged 20–49 years in 2015

| Variable | Age-adjusted prevalence [a] | | Prevalence ratio | 95% lognormal confidence interval | |
|---|---|---|---|---|---|
| | Filipino migrant women | Korean women | Filipino migrant women *versus* Korean women | Lower limit | Upper limit |
| Smoking status (current) | | | | | |
| 20–29 years | 3.3 | 6.3 | 0.52 | 0.36 | 0.77 |
| 30–39 years | 1.5 | 4.1 | 0.37 | 0.23 | 0.60 |
| 40–49 years | 1.3 | 3.6 | 0.36 | 0.14 | 0.97 |
| Any alcohol use | | | | | |
| 20–29 years | 19.0 | 59.7 | 0.32 | 0.27 | 0.37 |
| 30–39 years | 11.6 | 47.0 | 0.25 | 0.21 | 0.29 |
| 40–49 years | 9.1 | 37.6 | 0.24 | 0.17 | 0.35 |
| Physical inactivity | | | | | |
| 20–29 years | 59.1 | 25.4 | 2.33 | 2.13 | 2.56 |

| | | | | | |
|---|---|---|---|---|---|
| 30–39 years | 53.2 | 33.4 | 1.59 | 1.47 | 1.72 |
| 40–49 years | 45.6 | 32.6 | 1.39 | 1.19 | 1.64 |
| Low income level | | | | | |
| 20–29 years | 55.1 | 16.7 | 3.33 | 2.94 | 3.70 |
| 30–39 years | 41.0 | 15.6 | 2.63 | 2.38 | 2.94 |
| 40–49 years | 43.4 | 22.1 | 1.96 | 1.64 | 2.33 |
| Obesity | | | | | |
| 20–29 years | 14.8 | 12.0 | 1.23 | 1.03 | 1.49 |
| 30–39 years | 24.2 | 16.6 | 1.47 | 1.28 | 1.67 |
| 40–49 years | 41.4 | 23.5 | 1.75 | 1.47 | 2.08 |
| Hypertension | | | | | |
| 20–29 years | 1.1 | 1.3 | 0.88 | 0.45 | 1.72 |
| 30–39 years | 4.8 | 2.8 | 1.72 | 1.30 | 2.27 |
| 40–49 years | 19.4 | 9.7 | 2.00 | 1.54 | 2.56 |
| Diabetes | | | | | |

| | | | | | |
|---|---|---|---|---|---|
| 20–29 years | 1.2 | 0.7 | 1.72 | 0.88 | 3.33 |
| 30–39 years | 3.1 | 1.5 | 2.08 | 1.47 | 2.94 |
| 40–49 years | 3.9 | 3.4 | 1.14 | 0.65 | 2.00 |
| Hypercholesterolemia | | | | | |
| 20–29 years | 2.2 | 3.1 | 0.70 | 0.44 | 1.14 |
| 30–39 years | 4.3 | 6.0 | 0.72 | 0.54 | 0.97 |
| 40–49 years | 14.6 | 8.8 | 1.64 | 1.23 | 2.22 |

[a] Data are expressed as percentages.

**eTable 5**. Comparing age-adjusted health-related indicators between Filipino migrants and Koreans aged 20–49 years in 2015

| **Variable** | **Age-adjusted prevalence (95% CI)** | | **P-value** |
|---|---|---|---|
| | **Koreans** | **Filipino migrants** | |
| Men, n | 77,038 | 9,497 | |
| Smoking status (current) | 46.5 (45.9–47.1) | 23.3 (22.0–24.6) | <0.001 |
| Any alcohol use | 76.0 (75.3–76.8) | 53.6 (51.5–55.6) | <0.001 |
| Physical inactivity | 23.0 (22.6–23.4) | 48.3 (46.4–50.3) | <0.001 |
| Low income level | 8.8 (8.5–9.1) | 29.5 (28.2–30.9) | <0.001 |
| Obesity | 43.1 (42.5–43.6) | 40.0 (38.1–42.0) | 0.004 |
| Hypertension | 12.2 (11.9–12.4) | 10.0 (9.0–11.1) | <0.001 |
| Diabetes | 4.3 (4.2–4.5) | 3.9 (3.2–4.5) | 0.218 |
| Hypercholesterolemia | 10.3 (10.1–10.6) | 11.7 (10.6–12.7) | 0.010 |
| Women, n | 59,074 | 2,254 | |
| Smoking status (current) | 4.8 (4.5–5.0) | 2.1 (1.5–2.8) | <0.001 |
| Any alcohol use | 49.0 (48.3–49.6) | 13.6 (12.0–15.2) | <0.001 |
| Physical inactivity | 30.2 (29.7–30.7) | 53.2 (49.9–56.5) | <0.001 |
| Low income level | 17.9 (17.5–18.3) | 46.9 (43.8–50.0) | <0.001 |
| Obesity | 16.9 (16.5–17.2) | 25.7 (23.2–28.2) | <0.001 |
| Hypertension | 4.2 (4.1–4.4) | 7.7 (6.2–9.2) | <0.001 |
| Diabetes | 1.8 (1.7–1.9) | 2.6 (1.9–3.4) | 0.009 |
| Hypercholesterolemia | 5.7 (5.5–5.9) | 6.5 (5.2–7.9) | 0.246 |

Data are expressed as percentages; age standardization was performed using 10-year age bands; the World Standard Population was used as the standard population.

CI, confidence interval

### Online-Only References


1. Piao H, Yun JM, Shin A, Cho B. Longitudinal study of diabetic differences between international migrants and natives among the Asian population. *Biomol Ther (Seoul)*. 2020;28(1):110-118. doi: 10.4062/biomolther.2019.163.

2. Heng P. A Comparison of Cardiovascular Risk Factors among Asian Migrants and the Native Population in Korea. The Graduate School of Seoul National University; 2020.

3. Abubakar I, Aldridge RW, Devakumar D, et al. The UCL-Lancet Commission on Migration and Health: the health of a world on the move. *Lancet*. 2018;392(10164):2606-2654. doi: 10.1016/S0140-6736(18)32114-7.

4. Tziomalos K. Secondary hypertension: novel insights. *Curr Hypertens Rev*. 2020;16(1):11. doi: 10.2174/1573402115666190416161116.

5. Lee HY, Shin J, Kim GH, et al. 2018 Korean Society of Hypertension Guidelines for the management of hypertension: part II-diagnosis and treatment of hypertension. *Clin Hypertens*. 2019;25:20. doi: 10.1186/s40885-019-0124-x.
